# Outcomes After Percutaneous Tracheostomy in Patients with COVID-19: A Single-Center Series of 377 Cases

**DOI:** 10.1101/2022.12.28.22283971

**Authors:** Abdullah Alshukry, Abdulrazzaq Alhindi, Majdah Alzuabi, Seema Husain, Mohammad Tarakmeh, Shaima Al Qattan, Abdullah Al Bader, Ali Al Habib, Yaseen Ali, Fahd Al-Mulla, Hamad Ali, Mohammad Bu Abbas

## Abstract

**Introduction:** The COVID 19 pandemic was highlighted by a rise in hospital admissions secondary to respiratory decompensation. This was accompanied by an increase in ICU admissions, endotracheal intubation and mechanical ventilation. As a consequence, tracheostomies became essential in preventing complications of prolonged intubation and to facilitate weaning from sedation and mechanical ventilation. With the lack of international consensus on tracheostomy technique and optimal timing, we present our experience with 377 percutaneous tracheostomies performed on critically ill COVID 19 patients.

**Objective:** To report the outcomes of critically ill patients with COVID-19 who underwent percutaneous tracheostomy during a period of 24 months.

**Methods:** A retrospective single-center electronic chart review was performed on all ICU patients who underwent percutaneous tracheostomy after respiratory failure secondary to COVID-19 between March 2020 to March 2022.

**Results:** A total of 377 percutaneous tracheostomies were performed. The mean duration between intubation and percutaneous tracheostomy was 17.4 days (3-61). The study included 222 males (59%) and 155 females (41%).

The mean age of patients was 56.2 years (17-94), with a mean BMI was 31.3 (14-68).

The commonest comorbidities among patients were diabetes mellitus (50%) and hypertension (48%).

Complications were encountered in 85 cases (23%), with the commonest overall complication being minor bleeding.

203 patients (54%) were weaned from sedation. The mean duration between tracheostomy and weaning from sedation was 7.5 days (1 – 47 days). 156 patients (41%) were weaned from MV. The mean duration between tracheostomy and weaning from MV was 12.9 days (1 – 58 days). There was a total of 236 (63%) deaths reported during the period of this study.

No deaths were attributable to the surgical procedure.

**Conclusion:** Percutaneous tracheostomy can be safely performed in patients with COVID-19. With lack of conclusive objective data regarding the optimal timing for tracheostomy, we recommend that tracheostomy be performed as soon as possible after the 7^th^ day endotracheal intubation.

## INTRODUCTION

Since its emergence in Wuhan, China in December 2019, the new strain of the coronavirus, known as Severe Acute Respiratory Syndrome Coronavirus 2 (SARS-CoV-2), has rapidly spread around the world causing a coronavirus disease (COVID-19) pandemic with massive losses in financial, medical and human resources [1]. Kuwait officially reported its first positive case of SARS-CoV-2 on February 24, 2020, as passengers arrived on a plane from Iran [1]. As a response, the Kuwaiti Ministry of Health implemented strict measures to contain the pandemic, including the closure of schools and airports, curfews and prohibition of all public gatherings. Furthermore, the Ministry of Health designated a single center, Jaber Al Ahmad Al Sabah Hospital, as the country’s official COVID-19 center, thus receiving all confirmed cases of COVID-19 [2]. Nonetheless, the country was still severely affected by the consequences of this widespread disease, with 630,641 diagnosed cases and a total of 2,555 recorded deaths at the time of this report [3]. With the rapid rise in the number of positive cases of COVID-19, there was a parallel rise in Intensive Care Unit (ICU) bed occupation due to increasing numbers of patients requiring intubation secondary to the development of acute respiratory distress syndrome (ARDS) and other pulmonary complications of COVID-19. As many of these cases required prolonged intubation, tracheostomy was electively performed in order to facilitate pulmonary care, reduce the requirements of sedation and Mechanical Ventilation (MV), as well as reduce potential laryngotracheal complications associated with prolonged intubation. Due to the shortages in manpower and in aims of reducing the risk of viral contamination, a departmental decision was taken to perform tracheostomies using the percutaneous technique in preference to the classic open technique, whenever possible. This procedure would be performed at bedside, thus eliminating the need to transfer patients to and from the operating theater, requiring less medical and paramedical staff presence, therefore, reducing the risk of viral spread and cross contamination.

The aim of this article is to provide a descriptive analysis of our experience with 377 percutaneous tracheostomies that were performed in Jaber Al Ahmad Al Sabah Hospital between March 2020 to March 2022. To our knowledge, this is the world’s largest reported series of percutaneous tracheostomies from a single center.

## MATERIALS AND METHODS

### Study Design

A retrospective chart review of all patients admitted to Jaber Al Ahmad Al Sabah Hospital, Kuwait, with COVID-19 who required tracheostomy was carried out using the hospital’s electronic medical records database. All critically ill COVID-19 patients who were admitted to the ICU after intubation and mechanical ventilation and underwent tracheostomy between March 2020 and March 2022 were included. Recorded data included the following baseline characteristics: age, gender, body mass index (BMI), comorbidities (diabetes, dyslipidemia, hypertension, pre-existing cardiac disease (included coronary artery disease, heart failure, or a history of myocardial infarction), pre-existing pulmonary disease (included bronchial asthma, chronic obstructive pulmonary disease or pulmonary fibrosis), pre-existing renal disease (included nephropathies or renal failure), pre-existing coagulopathy, pre-existing malignancy, and any previous tracheal surgery. Furthermore, details on the duration of intubation prior to tracheostomy, time required for weaning from sedation and mechanical ventilation post tracheostomy, and complications were identified and recorded. Complications were classified as intra-operative, early (within 7 days of tracheostomy) or late (after 7 days of tracheostomy). Similarly, details were recorded if patients had ongoing dialysis or Extracorporeal Membrane Oxygenation (ECMO) during the procedure.

### Percutaneous Tracheostomy Protocol

Due to the lack of international consensus on the optimal timing for performing tracheostomy in COVID-19 patients, our department adopted a protocol of performing the procedure after a minimum of 7 days from endotracheal intubation. The mainstay of practice was to perform bedside percutaneous tracheostomy and reserve open tracheostomy for selected cases. Rarely, percutaneous tracheostomy was performed before the 7th day of intubation in selected patients in which prolonged intubation was anticipated at an earlier stage, such as patients with cerebrovascular accidents or other intra-cranial pathologies preventing potential rapid recovery of the level of consciousness.

The recommended MV settings for performing a percutaneous tracheostomy were:

- A Fraction of Inspired Oxygen (FIO_2_) of 70% or less, a Positive End-Expiratory Pressure (PEEP) of 12cm H_2_O or less, and a Respiratory Rate (RR) of less than 30 breaths per minute.

Laboratory investigations considered acceptable for percutaneous were as follows:

- Hemoglobin of 7 (g/dl) or above, platelet count of 50 (10^9^/L), International Normalized Ratio (INR) of 1.5 or less, Activated Partial Thromboplastin Time (APTT) of less than 50 seconds.

As for anti-coagulation treatment, which was routinely prescribed to COVID-19 patients in the ICU:

- Intravenous heparin infusions were stopped 4 hours prior to the procedure. Similarly, subcutaneous low molecular weight heparin was stopped 12 hours prior to the procedure. Any derangement in the coagulation profile secondary to anti-coagulation treatment was corrected prior to surgery.

The procedure was performed under bronchoscopy guidance at bedside in the ICU. A typical team would include 2 senior members of the otorhinolaryngology team – one guiding the procedure via bronchoscopy and the other performing the procedure. This was done in the presence of a member of the ICU anesthesia team, a respiratory technician and a staff nurse. Therefore, a maximum total of 5 members would be present at each procedure.

All staff members performing the procedure adhered to strict protective measures including the use of full personal protective equipment.

## RESULTS

Over a period of 24 months, between the dates of March 2020 – March 2022, a total of 377 percutaneous tracheostomy cases were performed on intubated and mechanically ventilated COVID-19 patients in the ICU.

The mean duration between intubation and percutaneous tracheostomy was 17.4 days (3-61).

The study included 222 males (59%) and 155 females (41%).

The mean age of patients was 56.2 years (17-94), with a mean BMI was 31.3 (14-68).

The commonest comorbidities among patients were diabetes mellitus (50%) and hypertension (48%). These were followed by dyslipidemia (18%), pre-existing pulmonary disease (16%), pre-existing cardiac disease (16%), and pre-existing renal disease (10%).

On the day of the procedure, 40 patients (8%) were on aspirin alone and 26 patients (5%) were on aspirin and Plavix. Due to contraindications imposed by underlying cardiac conditions, these medications were not stopped prior to surgery.

Furthermore, 109 patients (29%) had ongoing ECMO, and 86 patients (23%) had ongoing dialysis. These findings are summarized in Table 1.

**Table 1.** Clinical characteristics of COVID-19 patients included in the study.

| Variable | Number of Patients (%) (n = 377) |
| --- | --- |
| Age, mean ± SD (range) | 56.2 ± 15.3 (17 – 94) |
| Gender |  |
| Male | 222 (59%) |
| Female | 155 (41%) |
| BMI, mean ± SD (range) | 31.3 ± 7.1 (14.0 – 68) |
| Diabetes | 187 (50%) |
| Hypertension | 180 (48%) |
| Dyslipidemia | 63 (17%) |
| Pre-existing cardiac disease | 59 (16%) |
| Pre-existing pulmonary disease | 60 (16%) |
| Pre-existing renal disease | 38 (10%) |
| Malignancy | 12 (3%) |
| Smoker | 12 (3%) |
| Known coagulopathy | 8 (2%) |
| History of tracheal surgery | 0 (0%) |
| ECMO | 109 (29%) |
| Dialysis | 86 (23%) |

Complications were encountered in 85 cases (23%). These were classified as intra-operative, early or late.

Intra-operative complications were encountered in 23 cases (6%). The commonest intra-operative complication was minor bleeding which occurred in 18 cases (5%).

Early complications were encountered in 35 cases (9%). The commonest early complication was again minor bleeding which was encountered in 27 cases (7%). Furthermore, there were 2 cases of tracheo-esophageal fistula (0.5%) reported within 7 days of the procedure.

Late complications were encountered in 27 cases (7%). Similarly, the commonest late complication was minor bleeding which was reported in 9 cases (2%), followed by wound infection was which encountered in 8 cases (2%). Furthermore, there were 7 cases of tracheo-esophageal fistula (2%) and 1 case of tracheo-innominate fistula (0.2%), which underwent successful ligation. Full details on complications are summarized in Table 2.

**Table 2.**
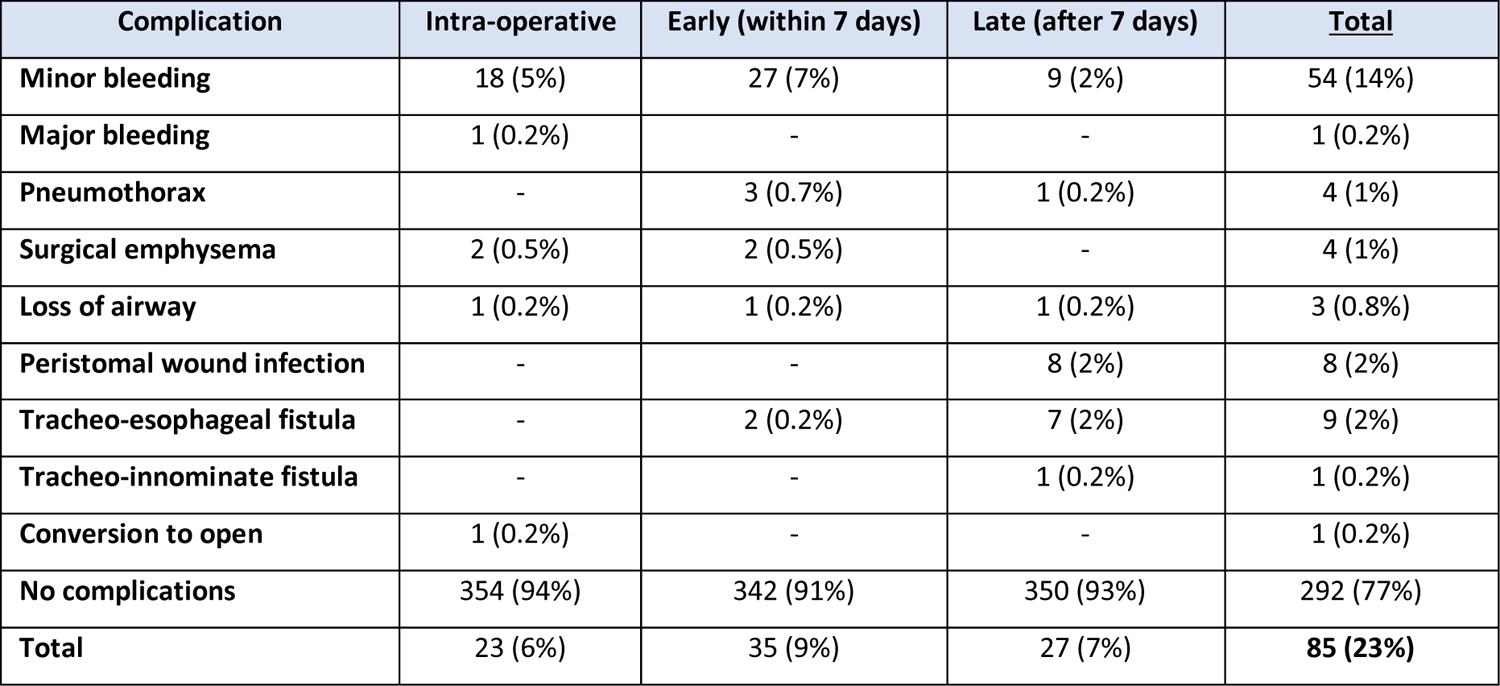
Rate of complications post percutaneous tracheostomy (n=377).

| Complication | Intra-operative | Early (within 7 days) | Late (after 7 days) | Total |
| --- | --- | --- | --- | --- |
| Minor bleeding | 18 (5%) | 27 (7%) | 9 (2%) | 54 (14%) |
| Major bleeding | 1 (0.2%) | - | - | 1 (0.2%) |
| Pneumothorax | - | 3 (0.7%) | 1 (0.2%) | 4 (1%) |
| Surgical emphysema | 2 (0.5%) | 2 (0.5%) | - | 4 (1%) |
| Loss of airway | 1 (0.2%) | 1 (0.2%) | 1 (0.2%) | 3 (0.8%) |
| Peristomal wound infection | - | - | 8 (2%) | 8 (2%) |
| Tracheo-esophageal fistula | - | 2 (0.2%) | 7 (2%) | 9 (2%) |
| Tracheo-innominate fistula | - | - | 1 (0.2%) | 1 (0.2%) |
| Conversion to open | 1 (0.2%) | - | - | 1 (0.2%) |
| No complications | 354 (94%) | 342 (91%) | 350 (93%) | 292 (77%) |
| **Total** | **23 (6%)** | **35 (9%)** | **27 (7%)** | **85 (23%)** |

Regarding the evolution of patients post tracheostomy, 203 patients (54%) were weaned from sedation. The mean duration between tracheostomy and weaning from sedation was 7.5 days (1 – 47 days). Furthermore, 156 patients (41%) were weaned from MV. The mean duration between tracheostomy and weaning from MV was 12.9 days (1 – 58 days). There was a total of 236 (63%) deaths reported during the period of this study. All deaths were related to COVID-19 complications and no deaths were attributable to complications of the surgical procedure. The remaining 141 patients (37%) were discharged, of which 53 patients (38%) were decannulated prior to discharge. The mean time from tracheostomy to decannulation was 36.5 days (6 – 82 days).

## DISCUSSION

The COVID-19 pandemic saw an exponential rise in hospital admissions for patients developing respiratory decompensation secondary to acute respiratory distress syndrome (ARDS). Not only would these patients develop severe symptoms requiring urgent in-patient medical attention, but the majority would require intubation and mechanical ventilation. According to the literature, the rates of intubation during the pandemic have been estimated at 10% to 12% of hospitalized patients and 58% of those admitted to the ICU [4 – 7]. Subsequently, the need for tracheostomies in the subset of patients requiring prolonged intubation and ventilation also saw a rise, with the main aims of the procedure being the facilitation of pulmonary care, the reduction of time required for weaning from sedation and MV, as well as the reduction of potential laryngotracheal complications. Unfortunately, the optimal timing and technique for performing a tracheostomy on COVID-19 patients was uncertain at the time and no international consensus was available [4, 8]. Different centers adopted different protocols, depending on the availability of the equipment and the experience of the staff. While some centers suggested positive outcomes whilst performing open tracheostomies, others advocated and advised for percutaneous tracheostomies.

In our center, the decision was taken to perform percutaneous tracheostomies for patients requiring intubation and MV beyond 7 days. The rationale behind this decision was the shorter duration of the procedure which can be performed at the patient’s bedside without the need for transfer to the operating theatre. Subsequently, a smaller number of medical and paramedical personnel were involved, thus reducing the risk of cross-contamination and viral spread.

Our series of 377 percutaneous tracheostomy cases over a period of 24 months represents the largest series reported in the literature. The mean duration of intubation to tracheostomy was 17.4 days in our series. This is comparable to the mean time reported by different studies, which ranges between 19 to 24 days [4, 9]. We believe that, ideally, tracheostomy should be performed as soon as possible after the 7^th^ day, however, due to the hemodynamic instability of patients with COVID-19 pneumonia, the delay is often longer, until a golden window is found.

In their report, Long et al. presented their outcomes of 101 tracheostomies performed for COVID-19 patients in their tertiary care center in New York City [4]. Of these 101 cases, 48 cases were percutaneous (48%) and 53 cases were open (52%). The authors report a total of 50 complications (50%), of which, the commonest complication was minor bleeding, which was encountered in 18 cases (18%) [4]. These complications were inclusive of both open and percutaneous techniques.

In our study, we report a lower rate of overall complications: 85 complications (23%). However, the rate of minor bleeding (14%) is comparable and was, likewise, the commonest complication encountered. All cases of minor bleeding were successfully managed by simple packing of the surgical site.

Long et al. reported 13 cases of peristomal infections, of which 9 (17%) were in the open tracheostomy group. The authors report a lower rate of wound infection in the percutaneous tracheostomy group - 4 cases (8%) [4]. In our study, we report a total number of 8 peristomal infections (2%). This would open the door for discussion as to whether percutaneous tracheostomy carries a lower risk of infection as less wound manipulation is required during the procedure, however, this is not within the scope of this article. Our findings are comparable to those reported by Chao et al. [9], in which wound infections were reported at 1.8%.

Alternatively, one of the major complications we report in this study is the detection of a total of 9 tracheo-esophageal fistulae (2%), of which 2 were detected within 7 days of tracheostomy. Although this is not directly caused by the surgical technique, it raises important points regarding the role of close monitoring of endotracheal tube and tracheostomy cuff pressure, especially in patients requiring prolonged intubation. In addition to the duration of intubation and the cuff pressure, these patients had multiple risk factors favoring the development of such complication. These factors included excessive motion of the tube during pulmonary care and patient proning, the presence of severe tracheo-broncho-pulmonary infections adding to the fragility of the trachea, as well as the administration of drugs that might potentially impede healing, such as steroids. Even though the use of high-volume low-pressure cuffs has reduced the incidence of tracheo-esophageal fistulae, long-term intubation still accounts for the majority of acquired cases because of either overinflation of the cuff or placement of a small tracheostomy tube requiring overinflation of the cuff to seal the airway [10 – 13].

In our report, the mean duration of weaning from sedation was 7.5 days after tracheostomy. There are very few articles in the literature reporting this outcome [4, 14, 15]. Long et al. reported a similar mean duration of about 7 days from tracheostomy to weaning from sedation [4]. In another report by Yeung et al., the mean duration to weaning from sedation was 2 days [14]. Similarly, Broderick et al. reported a mean duration to weaning of 24 hours in 10 patients post open tracheostomy [15]. Several reasons can account for these differences. First, the severity of the pulmonary infection can play a role in prolonging the duration of sedation. Second, the differences in the medications used for sedation were not accounted for in the analyses. Therefore, one must keep in consideration that certain drug regimen might interfere with the duration of time required for weaning.

Another valuable outcome measure is the mean duration of weaning from the MV after tracheostomy. In our report, the mean duration was 12.9 days after tracheostomy, which is comparable to that reported in the literature. Long et al. reported a mean duration for weaning from MV of 15 days after tracheostomy [4]. Similarly, Chao et al. reported a mean duration of 11.8 days [9]. In their study that included 148 percutaneous tracheostomies, Kwak et al. reported that 73% of their cases were weaned from MV after a mean duration of approximately 15 days following tracheostomy [16]. The results for this outcome seem similar across the different studies. Prolonged periods of sedation and mechanical ventilation result in a longer period of tube cuff inflation (endotracheal tube or tracheostomy tube), leading to complications that can range from simple infections to tracheomalacia to severe laryngo-tracheal complications such as stenosis or fistulae. These complications would ultimately compromise the patient’s prognosis. As a matter of fact, our data analysis suggests that the probability of survival decreases with a longer duration of sedation. For example, at 5 days of sedation, the probability of survival was approximately 75%, whereas at 28 days of sedation, the probability of survival was 50% (Figure 1). Similarly, patients requiring MV for less than 30 days had a survival probability of around 90%, and this probability decreases the longer MV is required. For example, patients requiring MV for longer than 45 days had a survival probability of less than 50% (Figure 2).

**Figure 1.**
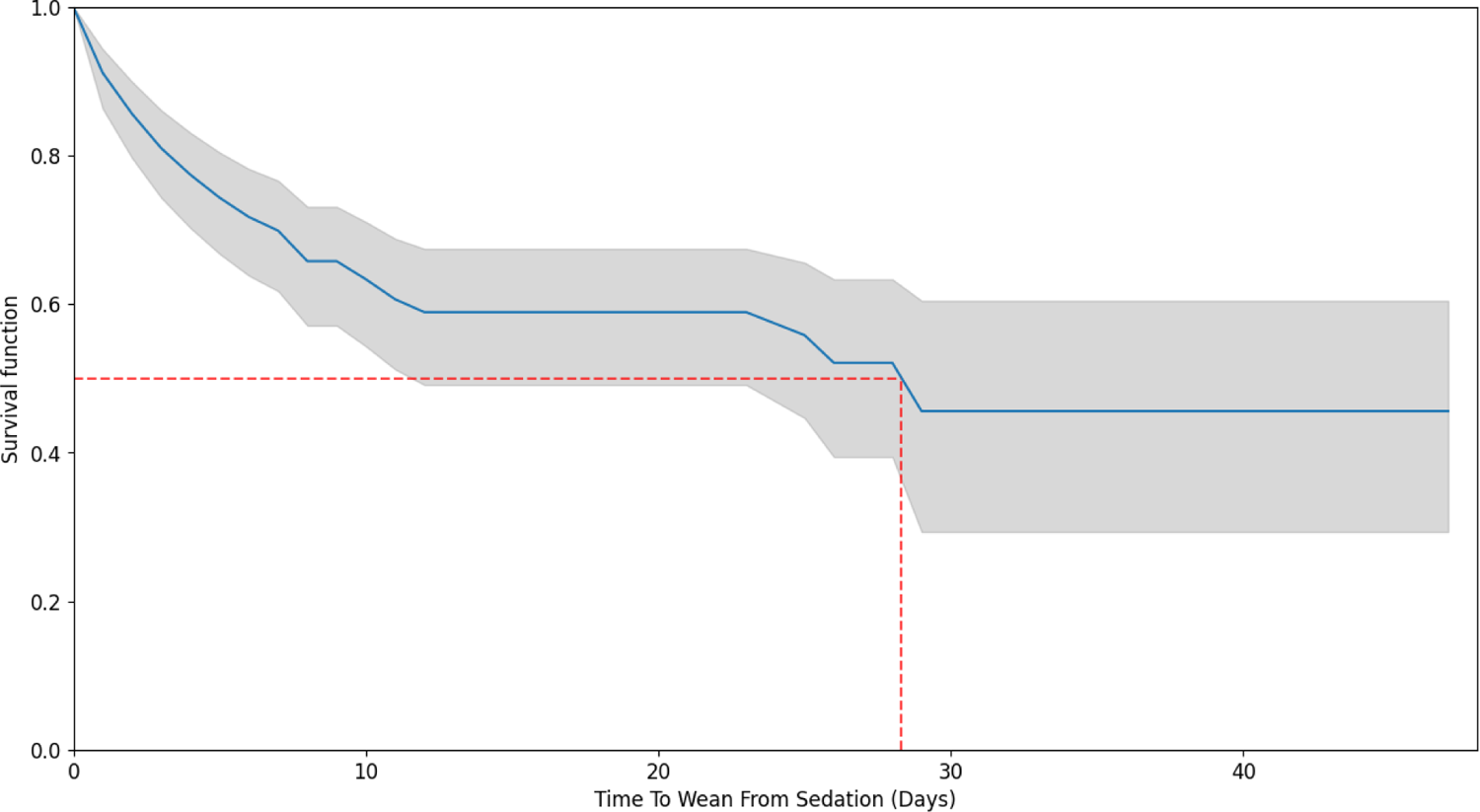
Kaplan Meir curve demonstrating the time to wean from sedation with the chance of survival associated with it.

**Figure 2.**
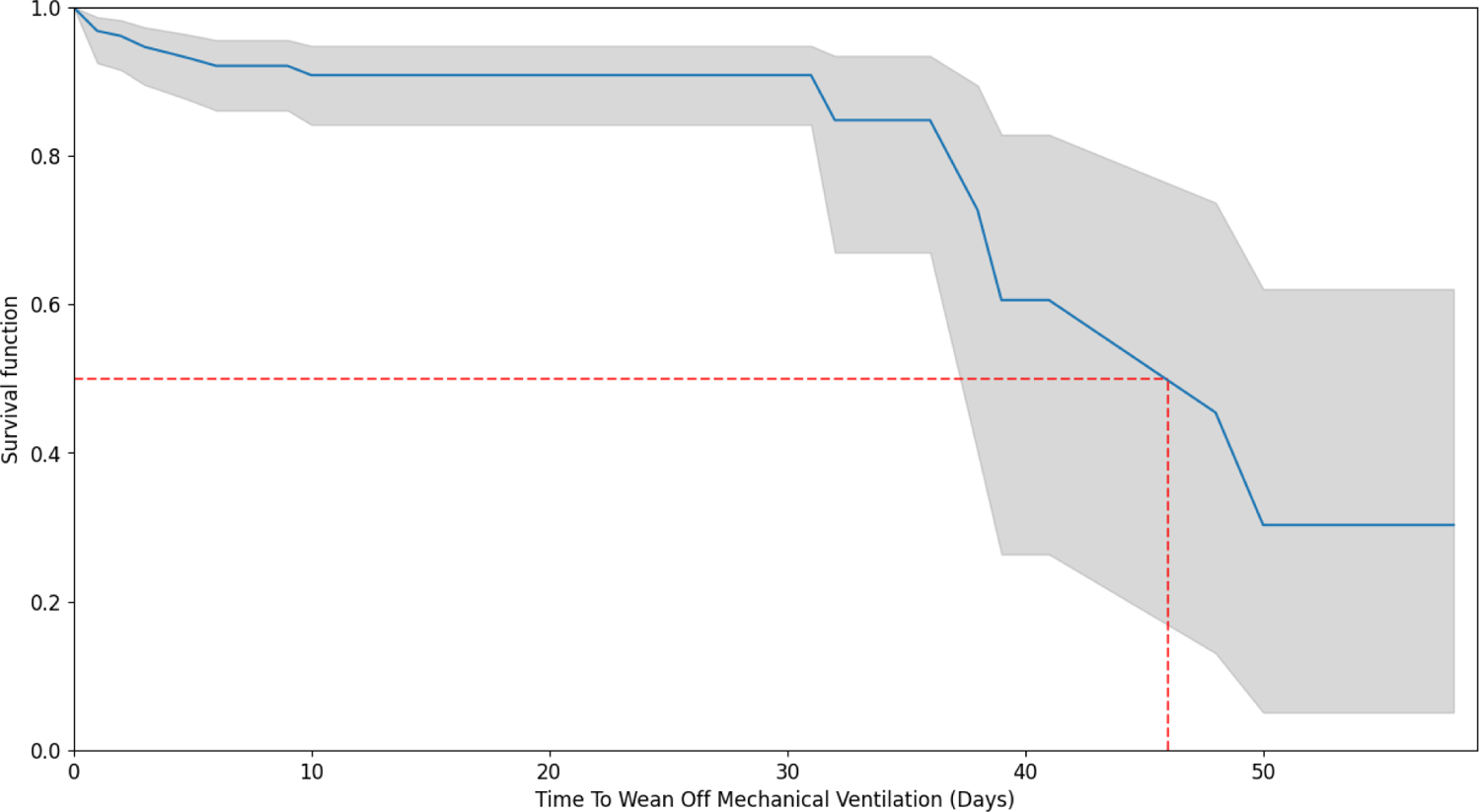
Kaplan Meir Curve demonstrating the time to wean from MV with the chance of survival associated with it.

Therefore, the results of our report suggest that percutaneous tracheostomy allowed for an overall reduction in the time required for weaning from sedation and mechanical ventilation.

The overall mortality rate in our study population was 63% (236/377). All deaths were secondary to COVID-19 complications with the commonest cause of death being cardiac arrest (37%). This high mortality rate reflects the severity of the disease in the patients included. Furthermore, the role of co-morbidities on survival must be considered. In our study, approximately 50% of patients suffered from diabetes or hypertension. Furthermore, 30% of patients were above the age of 65. The role of risk ratios was used to compare the relative risk between comorbidities among the study population. Of which, hypertension (RR=1.30, p=0.0015) and age > 65 (RR=1.24, p=0.0091) were found to have a significant association with increased risk for death. In the available literature at the time, intubated COVID-19 patients requiring MV exhibited a very poor prognosis with a very high risk of mortality. Reported mortality rates for mechanically ventilated COVID-19 patients ranged from 40% to 68% and this rate was higher in patients requiring additional supportive measures [17 – 19]. In our study, 29% of patients who required tracheostomy were on ECMO, and 23% were on dialysis. Furthermore, the role of ethnicity in COVID-19 outcome disparities should not be omitted. Kuwait has a unique demographic profile with more than 70% of population consisting of non-Kuwaitis [20]. With the majority of non-Kuwaitis being of South Asian nationalities, this ethnic group was specifically reported to have a worse prognosis and outcome when compared to patients of Arab ethnicity [21].

## CONCLUSION

Our analysis supports the growing body of literature suggesting that percutaneous tracheostomy can be safely performed in patients with COVID-19. We believe that in order to avoid complications and facilitate weaning from sedation and mechanical ventilation, and the risks associated with this, tracheostomy should be performed as soon as possible after the 7^th^ day of endotracheal intubation. However, in the absence of conclusive objective data, the question of optimal timing remains opens for debate.

## LIMITATIONS

Due to the nature of the pandemic and the departmental decision to perform percutaneous tracheostomies as the mainstay of practice, our study did not include an arm of patients who underwent open tracheostomy. This would have allowed a direct comparison of the two techniques in terms of outcomes.

Furthermore, patients in this study were followed up until one of two outcomes is reached: death or discharge. Therefore, long term outcomes such as speech and swallowing post decannulation are outside the scope of this report. Similarly, since many patients were discharged prior to decannulation, information on tracheostomy status of these patients were not included.

Finally, although, clinically, no members of staff performing the percutaneous tracheostomies developed COVID-19 during the period of this study, objective PCR testing was not performed. This would have provided an additional argument for the safety index of percutaneous tracheostomies.

## Data Availability

All data produced in the present work are contained in the manuscript

